# Integration of Hybrid Model Diabetes Self-Management Education and Support at Primary Health Care During COVID-19 Pandemic: Protocol Paper of DIAJAPRI Health Coaching Study

**DOI:** 10.1101/2022.12.18.22283494

**Authors:** Em Yunir, Syahidatul Wafa, Dicky L. Tahapary, Lusiani Rusdi, Yeni D. Lestari, Pringgodigdo Nugroho, Annisa P. Nachrowi, Anandhara I. Khumaedi, Tri J.E. Tarigan, Simon Salim, Gitalisa A. Adriono, Arif Mansjoer, Sarwono Waspadji, Imam Subekti, Dante S. Harbuwono, Suharko Soebardi, Budiman Darmowidjojo, Dyah Purnamasari, Wismandari Wisnu, Farid Kurniawan, Martha Rosana, Ardy Wildan, Eunike V. Christabel, Tika Pradnjaparamita, Nida Amalina, Endang S. Wahyuningsih, SW Novita, Fitri Damayanti, Vinny Vanessa, Idrus Alwi, TLH Dwi Oktavia, Ratna Sitompul, Pradana Soewondo

## Abstract

**Introduction:** COVID -19 pandemic has threatened the optimal achievement on type-2 diabetes mellitus (T2DM) target in primary health care (PHC), due to our priority in COVID-19 management, limited access of patients to PHC and their lifestyle changes as the impact of social restrictions. Therefore, the empowerment of capability of patients on diabetes self-care is required through optimal education and support. The use of telehealth in T2DM management has benefits on improving outcomes of patients. We aim to assess the role of telehealth diabetes self-management education (DSME) versus hybrid (telehealth and face-to-face method) diabetes self-management education and support (DSMES) to improve T2DM outcomes in PHC during COVID-19 pandemic.

**Methods and analysis:** This study is an open label randomized-controlled trial that will be conducted in 10 PHCs in Jakarta, Indonesia, involving patients with T2DM. Subjects are classified into 2 groups: DSME group and DSMES group. Intervention will be given every 2 weeks. DSME group will receive 1 educational video every 2 weeks discussing topics about diabetes self-management, while DSMES group will receive 1 educational video and undergo 1 coaching session every 2 weeks. All interventions will be conducted by trained health workers of PHC, who are physicians, nurses, and nutritionists. Our primary outcome is the change of HbA1C level and our secondary outcomes are the changes of nutritional intake, physical activity, quality of life, anthropometric parameter, fasting blood glucose, lipid profile, inflammatory markers, and progression of diabetes complications at 3 and 6 months after intervention compare to the baseline.

**Ethics and dissemination:** This study protocol has been approved by the Health Research Ethics Committee University of Indonesia. Subjects agree to participate will be given written informed consent prior to data collection. Findings from this study will be published in peer-reviewed journals and presented at conferences.

**Trial Registration:** http://www.clinicalstrials.gov with identifier number NCT05090488.

**Summary:** *Strengths and limitations of the study:*

- This study evaluates the role of hybrid DSMES, which is useful in areas with limited access or on lockdowns.
- This study will evaluates the implementation of hybrid DSMES, its benefits, difficulties, and obstacles.
- We uses validated questionnaire instruments and routinely collected clinical data.
- Because all of our interventions will be conducted by PHCs’ health workers, our results depend on the ability and adherence of PHCs’ health workers.

## Background

Type-2 diabetes mellitus (T2DM) remains a global burden. In 2019, the prevalence of T2DM in adult population was about 463 million and is predicted to become 700 million in 2045 (1). Indonesia is one of the top ten countries with highest number of people with T2DM (2). Based on 2018 Indonesian National Health Research, T2DM is one of the major national health problem with rising prevalence from 6.9% in 2013 to 10.9% in 2018 (3). The progression of T2DM leads not only to a debilitating micro and macrovascular complications that decrease patients’ quality of life and life expectancy, but also increases health expenditure (1).

Even more, T2DM epidemic is worsened by the emergence of COVID-19 pandemic (4). COVID-19 pandemic has provided barriers for patients with diabetes to access health care either for routine or emergency medical care. The regulation of large scale of social restrictions (LSSR) to limit COVID-19 dispersion rendered patients to mostly stay at home, thereby increasing unhealthy lifestyle risk factors such as poor diet, lack of physical activity, feeling of isolation/loneliness, and psychological stress. COVID-19 pandemic also threatens the comprehensive diabetes care, which requires regular patient-provider interactions for education, prescriptions, and assessment of complication and patient’s well-being. Previous study in Indonesia reported that LSSR ultimately raises the incidence of diabetes complications (5). This fact highlights the importance of patient’s self-management in order to achieve optimal target, particularly during COVID-19 pandemic.

Diabetes self-management education and support (DSME/S) provides the foundation to help patient with diabetes to have good knowledge and ability of diabetes self-care. DSME is a process in which health care team facilitates patients with knowledge and skill necessary to manage themselves, whilst DSMS refers to the support required to integrate and sustain expected behaviors (6). DSMES model is specifically designed to elaborate the biological, psychological, social, cultural, and economic aspects of each individual with T2DM and enable them to navigate their own daily activities. Our previous hospital-based study reported that DSMES is superior than DSME (both using face-to-face method) in lowering A1C level in patients with diabetes, although the benefit on body weight reduction and improvement of blood pressure and lipid profile was not satisfying (7). However, another studies have shown that DSMES effectively improve patient’s knowledge, self-care behaviors, glycemic control (8), and quality of life (9), and finally reduce of all-cause mortality risk (10) and health cost (11). The implementation of DSMES in primary health care (PHC) as the first line health center closest and most readily accessible by patients should become a priority.

PHCs play an important role in preventive and promotive measures. At PHCs, physicians and health workers have routinely performed T2DM standard of care, including education to promote lifestyle changes and medication adherence. However, as the global concern for COVID-19 has put majority focus on hospital care, the continuity care for chronic disease such as T2DM is considered overlooked. Hence, the integration of diabetes self-support in the form of DSMES into T2DM standard of care in PHC should be encouraged particularly during COVID-19 pandemic (6).

Unfortunately, implementation of DSMES during COVID-19 pandemic is another challenge. Health workers capacity is limited during pandemic because the major priority was to contain the spread of COVID-19. Access to health care is also limited due to LSSR and fear to visit health centers. Therefore, integration of DSMES approach with telemedicine during COVID-19 pandemic setting is required in order to optimize outcomes (12). Several studies had shown that usage of telehealth in T2DM patients significantly improve not only glycemic control (13–15), but also body weight, blood pressure, lipid profile (16,17), and ultimately, quality of life (18). This study is an open-label randomized controlled trial (RCT) aim to assess the role of telehealth DSME versus hybrid DSMES (combination of face-to-face and telemedicine approach) in T2DM patients at PHC during COVID-19 pandemic, namely DIAJAPRI (Diabetes in Jakarta Primary Care) Health Coaching Study. We address the changes of glycemic control, behavior (nutritional intake and physical activity), macro and microvascular complications, metabolic and inflammatory parameters, and quality of life.

## Methods

### Study area

This study will be conducted in Jakarta, Indonesia. Jakarta is the capital city of Indonesia and is the sixth most populated city in Indonesia with 10.5 million people (19). It is the largest city in Southeast Asia by population number, one of the cities with fastest growing economies with urban population characteristics (20). Based on National Health Research 2018, Jakarta is the province with highest prevalence of diagnosed diabetes patients, rising from 2.5% in 2013 to 3.4% in 2018 (3).

### Patient and Public Involvement

We collaborate with Provincial Health Agency and PHC health workers when we design and conduct this study. We discuss and modify our study protocol according to input from PHC health workers. They will also recruit the patients involved in this study, according to the inclusion and exclusion criteria. Finally, PHC health workers will be trained and provide DSME and DSMES sessions for patients under our supervision.

### Study design

This study is an open label randomized-controlled trial that will be conducted in 10 district PHCs Jakarta, Indonesia. Each PHC recruits 20 patients with T2DM consecutively who meet the inclusion and exclusion criteria. We will randomly classify subjects into 2 groups: DSME group and DSMES group, based on 4:1 block randomization technique aided by computer software (Random Allocation Software). Intervention will be given every 2 weeks. DSME group will receive 1 educational video every 2 weeks discussing topics about diabetes self-management, while DSMES group will receive 1 educational video and undergo 1 hybrid coaching session every 2 weeks. Coaching intervention consists of an initial face-to-face session, followed by 5 hybrid sessions that combine telemedicine or face-to-face approach. Measurements will be collected at baseline, 3 months, and 6 months after intervention. Study protocol can be seen in Figure 1.

**Figure 1.**
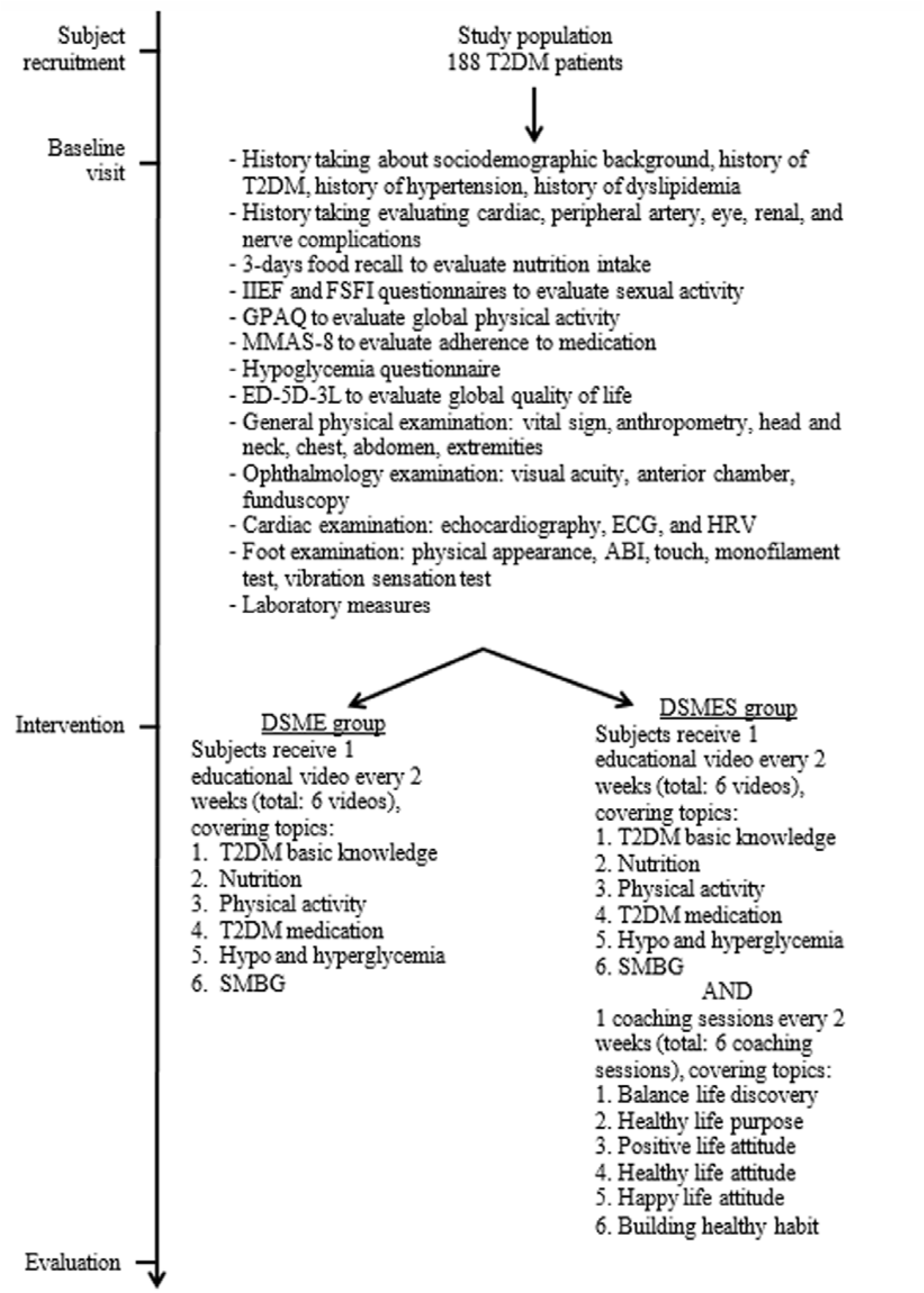
Description of study protocol

All interventions will be conducted by PHC’s health workers as the representative of non-communicable disease (NCD) clinic team, consisting of 1 physician, 1 nurse, and 1 nutritionist. Health workers must join an online health coaching workshop and fulfill minimum coaching practice hours to become a certified coach. Details will be explained below. Each 1 health workers will be responsible to educate and/or coach 3-4 T2DM subjects, who will be randomly assigned by researchers.

### Inclusion and exclusion criteria

Each participating PHC consecutively selects patients aged ≥ 18 years old, either male or female, and have been diagnosed with T2DM according to ADA criteria: (1) Fasting plasma glucose (FPG) ≥ 126 mg/dl, or (2) 2-hours plasma glucose (2-h PG) ≥ 200 mg/dl during oral glucose tolerance test (OGTT), or (3) A1C ≥ 6.5%, or (4) patient with classic symptoms of hyperglycemia and random plasma glucose (RPG) ≥ 200 mg/dl. Recruited subjects should sign an informed consent prior to the study. Subjects unable to communicate well, such as patients with cognitive problems (such as dementia), hearing or sight problems, and disability to live independently on daily basis are excluded from the study.

### Intervention

#### CARE coaching model training

Prior to DSMES sessions, health workers, who are physicians, nurses, and nutritionists, from 10 PHCs will be trained. Training was based on CARE (Clarity, Awakening, Resolution, and Empowerment) coaching model, according to Positive Psychology Coaching model (21), which focus on developing solution-focused thinking. CARE coaching model describes the steps that must be achieved in each coaching session. Coaching sessions use patient-centered approach to support patient to set their goals and the step needed to achieve goals. The focus of coaching is to develop problem solving strategy and focus on goals achievement.

In the Clarity step, coaches will help patients to set a specific goal aimed from each session. Coaches will give several open-ended questions to trigger patients to set their own goals. These goals should be specific, clear, relevant, achievable, and measurable. Patients will also set a certain period of time when they should have achieved their goals. In the Awakening step, coaches will help patients to raise self-awareness about their values, capacities, and qualities. In the Resolution step, coaches will help patients to take decision and make priority scale to overcome their obstacles. In the Empowerment step, coaches will close the coaching session by empowering patients to be focus and responsible to their goals.

This CARE coaching model training will be given for 4 days, 4 hours per day each. Topics covered in the training sessions are (1) basic theory of T2DM, (2) introduction of coaching theory (group clarity, definition of coaching, basic coaching principles, medical practitioner as a coach, definition of health coaching, roles of health coach, and scope of health coaching), (3) CARE coaching theory, (4) basic coaching theory, (5) clarity theory, (6) awakening theory, (7) resolution theory, (8) commitment theory, and (9) structured health counseling and CARE coaching model. Each of the session consists of coaching demo, debriefing, and self-exercise.

Every coach should continuously apply coaching technique in their clinical practice and record the coaching practice in a logbook. The coaching portfolio should be submitted to the trainer and then will be discussed in a separated groups supervised by a certified coach.

#### DSMES intervention

Recruited subjects will be consecutively divided into 2 arms: DSME group and DSMES group. Subjects in the DSME group will receive 1 educational video made by researchers every 2 weeks for a total of 6 sessions. Each video run for about 20 minutes, discussing about T2DM and its management. Topics covered are (1) basic knowledge about T2DM, (2) nutrition and diet for T2DM patients, (3) physical activity for T2DM patients, (4) T2DM medications, (5) hypoglycemia and hyperglycemia, and (6) self-monitoring of blood glucose (SMBG). These educational videos will be broadcasted online to the T2DM subjects. Meanwhile, subjects in the DSMES group will receive the same educational video every 2 weeks for a total of 6 sessions, with an additional of 6 coaching sessions with hybrid model. To enhance subjects’ understanding about the topic, the person in charge (PIC) from each PHC will open a question-and-answer session with all subjects. The PIC may also give several questions regarding to the topics covered in the video to generate curiosity and group discussion.

Coaching sessions for subjects in the DSMES group will be given as 1 coaching session every 2 weeks, for a total of 6 sessions. These coaching sessions are based on CARE coaching model. In every coaching session, 1 coach and 1 subject will meet for 45-60 minutes and discuss 1 specific topic. Topics covered in these 6 sessions are (1) balance life discovery, (2) healthy life purpose, (3) positive life attitude, (4) healthy life attitude, (5) happy life attitude, and (6) building healthy habit.

Research team members will supervise the whole process using written logbook and online meeting every 2 weeks. There are 3 different logbooks, distributed for DSME subjects, DSMES subjects, and coaches. Logbook for DSME subjects consist of essay questions regarding topics discussed in the video every 2 weeks period. These questions will assess subjects’ knowledge and understanding about the video. During the 2-weeks period, subjects have to show their logbook to the PIC in the PHC and ask for their signature. The PIC will make sure if subjects’ already watch the video and understand the topics discussed. Similarly, in the logbook for coaches, the PIC will write their subjective assessment about subjects’ understanding regarding the topics discussed. Logbook for DSMES subjects also consist of essay questions similar to that of the logbook for DSME subjects, however they will have additional pages for coaching sessions. Each of the targets and topics discussed with their coach in every coaching session should be written down in the logbook and signed by their coach. Similarly, in the logbook for coaches, each coach will also evaluate their subjects during every coaching session. Every 2 weeks, in the end of the 2-weeks period before moving to another topics, online meeting with research team members will be held to evaluate and record the progress, difficulties, and obstacles encountered by each PHCs.

#### Outcomes

The main outcome of this study is the change of HbA1C level in patients with T2DM in DSMES compared to DSME group at 3 and 6 months after intervention. Secondary outcomes are changes of mean calorie intake, global physical activity, quality of life, anthropometric parameter (body mass index, waist circumference, body fat), fasting blood glucose, serum lipid profile (total cholesterol, LDL-cholesterol, HDL-cholesterol, and triglycerides), inflammatory marker (c-reactive protein), and progression of diabetes complications (eye, cardiac, kidney, peripheral artery, neuropathy, and sexual function). All secondary outcomes will also be compared among DSMES versus DSME group at 3 and 6 months after intervention compared to baseline. The detailed explanations of outcomes measurement are explained below.

#### Sample size

Sample size is calculated using test for difference in 2 independent arms. According to the calculation, minimum number of subjects for each arm was 90 subjects. Assuming that 10% of the subjects may be lost to follow up, we decide to recruit 200 subjects, in which 100 subjects will be classified as DSME group and 100 subjects will be classified as DSMES group. We use 5% significance level and a power of 80%.

#### Baseline visit interview

Upon acceptance into the study, subjects will be interviewed by the physicians in each PHC, according to the questionnaires provided. First questionnaire covers sociodemographic information, such as gender, date of birth, residence, ethnicity, religion, marital status, occupation, education, income, health insurance, and smoking habits), history of T2DM (duration of T2DM, family history of T2DM, and history T2DM medication), history of hypertension (duration of hypertension and history anti-hypertensive therapy), and history of hypercholesterolemia (duration of hypercholesterolemia and history of cholesterol medication).

Second questionnaire evaluates subjective complaints suggestive of T2DM complications, evaluating symptoms of macroangiopathy (cardiac and peripheral artery complications) and microangiopathy (eye, renal, and nervous complications). Questions about cardiac complications include: chest pain, dyspnea on effort, swollen feet, palpitations, and history of coronary interventions. Questions about peripheral artery complications include: pain, paresthesia, and tingling on right, left, or both feet. Questions about retinopathy include: blurred vision, history of cataract, and history of eye laser. Questions about nephropathy include: foamy urine, decreasing urine output, and history of dialysis.

Third questionnaire evaluates nutrition intake, sexual activity, family support, physical activity, awareness to hypoglycemia event, and quality of life. Nutrition intake will be assessed using 3-days food recall prior to baseline visit. Food list will be analyzed using NutriSurvey (2007) application from http://nutrisurvey.de for Indonesia. Through this application we generate average carbohydrate, protein, and fat intake (in gram and kcal) and its percentage fulfillment according to daily average requirement.

Sexual activity will be assessed using International Index of Erectile Function (IIEF) questionnaire (22) for male subjects and Female Sexual Function Index (FSFI) questionnaire (23) for female subjects. IIEF questionnaire consists of 5 domains (erectile function, orgasmic function, sexual desire, intercourse satisfaction, and overall satisfaction); however, in this study we only assess the first domain, which is erectile function. This domain consists of 6 questions, each score 0-5 (0 for no sexual activity and 5 for always or almost always satisfied). Subjects score < 14 out of 30 points are regarded to have erectile dysfunction. FSFI questionnaire consists of 6 domains (desire, arousal, lubrication, orgasm, satisfaction, and pain) and a total of 19 questions, each score 0-5 (0 for no sexual activity and 5 for always or almost always satisfied). Point from each domain is multiplied with constant factor (0.6 for desire, 0.3 for arousal and lubrication, and 0.4 for orgasm, satisfaction, and pain). Subjects score ≤ 26.55 out of 36.0 are regarded to have sexual dysfunction.

Global physical activity is assessed using Global Physical Activity Questionnaire (GPAQ) by WHO (24). This questionnaire consists of 4 domains (activity at work, travel to and from places, recreational activities, and sedentary behavior) and a total of 16 questions. This questionnaire measures total minutes of moderate-to-vigorous physical activity (MVPA) and its metabolic equivalent units (MET). Awareness to hypoglycemia events will be assessed using hypoglycemia questionnaire from Hypoglycemia Health Association of Australia (25). This tool consists of 10 questions covering symptoms of hypoglycemia. Each question score 0 for never, 1 for rarely, 2 for occasionally, and 3 for usually. Subjects with total score <8 are unlikely to have hypoglycemic disease, score 8-15 are possibly having hypoglycemic disease, and score >15 are certainly having hypoglycemic disease. Global quality of life will be assessed using EuroQol-5-dimension-3-level (EQ-5D-3L) questionnaire (26). This tool consists of 5 domains (mobility, self-care, usual activities, pain/discomfort, and anxiety/depression). Each dimension has 3 levels: 1 point for no problem, 2 points for some problems, and 3 for extreme problems. The maximum total score is 5, which indicates best health state, while the minimum total score is 15, which indicates worst health state.

#### Baseline visit examination General physical examination

General physical examination will be conducted to evaluate the general condition of the subject. This includes vital signs, anthropometric measurements, head and neck, chest, abdomen, and extremities examination. Vital signs include: blood pressure, pulse rate, respiratory rate, body temperature, and peripheral oxygen saturation. Blood pressure is measured in non-dominating arm, sitting position, after 15 minutes rest using digital sphygmomanometer (HEM-7200, Omron Healthcare Co, Kyoto, Japan). Sphygmomanometer is calibrated using Riester nova-presameter desk mercury sphygmomanometer (Gerhard Glufke Rudolf Riester Gmbh, Germany) prior to usage. Pulse rate is measured manually by palpating brachial artery in non-dominating arm for 1 minute. Pulse strength, rate, and rhythm are reported. Respiratory rate is measured manually for 1 minute. Body temperature is measured using non-contact medical infrared thermometer (HCO Wready Care Infrared Non-contact Thermometer WDKL-EWQ-001, Hunan Weiding Information Technology Co, Ltd, China). Peripheral oxygen saturation is measured using pulse oximetry (FOX-3 Elitech pulse oximetry, Elitech Technovision, Indonesia).

Anthropometric measurements include: body weight, body height, waist circumference, and body fat composition. Measurements are obtained according to National Heart, Lung, and Blood Institute (NHLBI) guidelines. Body weight is measured using a flat scale (SECA Model 876, Seca Gmbh Co, Hamburg, Germany) to the nearest 0.1 kilograms. Body height is measured using portable stadiometer (SECA Model 213, Seca Gmbh Co, Hamburg, Germany) to the nearest 0.1 centimeter. Using body weight and body height, body mass index is calculated. Waist circumference is measured using circumference measuring tape (SECA Model 201, Seca Gmbh Co, Hamburg, Germany). Body fat composition (fat percentage, fat mass, muscle mass, bone mass, basal metabolic rate, metabolic age, water ratio, and visceral fat) is measured using TANITA body impedance analyzer (TBF-300A, Tanita Corp, Tokyo, Japan).

Head and neck examination includes: conjunctiva (pale/normal), sclera (icteric/not), and thyroid (palpable/not). Chest examination includes: heart (regularity of heart rate, presence of gallop, and presence of murmur) and lung (presence of rhonchi and wheezing). Auscultation uses stethoscope (3M Littmann Classic II SE Stethoscope, 3M, Minnesota, USA). Abdomen examination includes: ascites, hepatomegaly, splenomegaly, and kidney ballottement. Extremities examination includes: capillary refill time and edema.

#### Detailed examination

Detailed clinical examination is conducted to evaluate signs of T2DM complications, which are eye, cardiac, peripheral artery, and nerve complications.

#### Ophthalmology examination

Ophthalmology examinations are performed by an ophthalmologist including visual acuity, segment anterior and posterior. Presenting visual acuity(VA) measurement is performed at 2 meters by using PEEK® acuity chart application and pinhole is used when VA worst than 6/6. This application can measure visual acuity from no light perception to 6/6. Subjects using spectacles will be asked to use their glasses throughout the VA measurement. Visual acuity is classified according to the classification by World Health Organization (27). Patients are categorized as blind if presenting VA is lower than 3/60 in the better eye, severe visual impairment (SVI) if presenting VA is lower than 6/60 to 3/60, moderate visual impairment (MVI) if presenting VA is lower than 6/18 to 6/60, mild visual impairment if presenting VA is lower than 6/12 to 6/18, and categorized as normal if presenting VA is equal or higher than 6/12.

Anterior segment examination includes eyeball position, eyeball movement, palpebra, conjunctiva, cornea, anterior chamber depth, iris, and pupil condition using loupe and penlight. Lens haziness is evaluated using shadow test. Subjects are examined for pathologic conditions, including cataract, refraction error, or glaucoma, etc.

Posterior segment examination is conducted by taking ocular fundus photography using Topcon TRC-NW400 fundus camera. Fundus images is evaluated and graded for diabetic retinopathy (DR) by a vitreoretinal specialist. Subjects will be classified into no (DR), mild non-proliferative diabetic retinopathy (NPDR), moderate NPDR, severe NPDR, and proliferative diabetic retinopathy (PDR) using International Classification of Diabetic Retinopathy and Diabetic Macular Edema (28).

Subjects are classified as mild NPDR if only microaneurysm presents; moderate NPDR if dot-blot hemorrhages, cotton wool spots, and hard exudates present; severe NPDR if intraretinal hemorrhage (more than or equal to 20 in each of four quadrants), venous beading in at least 2 quadrants, or intra-retinal microvascular abnormalities (IRMA) in at least 1 quadrant present; and PDR if neovascularization of optic disc or elsewhere, vitreous hemorrhage, or preretinal hemorrhage present (28). DR grading will be assessed for each eye. Subjects with different stage of DR in each eye will be categorized into the most severe condition. Fundus image with questionable results is classified into ‘ungradable’.

#### Cardiac examination

Cardiac evaluations conducted are echocardiography, electrocardiography (ECG), and heart rate variability (HRV) examination. Echocardiography at rest is performed using Philips CX-50 and transducer sector S5-1. Linear measurements of the left ventricle (LV) and its walls are performed through 2D guided M-mode approach or 2D echocardiographic imaging in parasternal long-axis, as recommended by American Society of Echocardiography. LV mass index is calculated using standard formula and corrected by body surface area (BSA) (29). We define left ventricle hypertrophy (LVH) if LV mass ratio to BSA is >115 g/m^2^ in men or >95 g/m^2^ in women (30). LV systolic function is calculated from ejection fraction (four-chamber view) and global longitudinal strain (GLS). Measurement of the left atrial (LA) volume is performed through biplane area calculation. LA volume is indexed according to BSA. LA enlargement is defined as LA volume ratio to BSA more than 34 ml/m^2^. Right ventricle (RV) systolic function is evaluated using tricuspid annular plane systolic excursion (TAPSE), measured by M-mode echocardiography with cursor optimally aligned along the direction of tricuspid lateral annulus in the apical four-chamber view (29). LV diastolic function is evaluated according to the algorithm recommended by American Society of Echocardiography in 2016 (31).

ECG and HRV examination are done using ECG Holter Type 1303, Vasomedical BIOX ™. Patients lie down, and 12 lead-ECG recording is recorded for 10 minutes. The recording is processed using ARCS Series ECG and ABP Analysis and Reporting Software Ver 8.0.7 to produce 10-minutes HRV result and a regular ECG strip.

#### Foot examination

Foot examination is done to assess diabetic foot, amputation, peripheral artery disease (PAD), and diabetic peripheral neuropathy. Examination is conducted in both right and left feet. Diabetic foot is evaluated visually by presence of hair thinning, deformity of the bone, edema, hyperpigmentation, atrophy, callus, nail abnormalities, dry skin, and healed wounds (32). Amputation is the resection of a segment of a limb through a bone or joint. Major amputation is any resection proximal to the ankle. Minor amputation is any resection through or distal to the ankle. Amputation is also evaluated visually (32). The subjects’ feet will be photographed and confirmed by an endocrinologist.

PAD is an obstructive atherosclerotic vascular disease with clinical symptoms, signs, or abnormalities on either non-invasive or invasive vascular assessment, resulting in disturbed or impaired circulation in one or more extremities (32). PAD is diagnosed through ankle brachial index (ABI) measurement (33). Pulse strength and blood pressure is assessed in right and left brachial artery, dorsal pedis artery, and posterior tibial artery. Highest brachial and ankle systolic blood pressure is used to calculate ABI. ABI will be classified according to the guideline from the Indonesian Society of Endocrinologist (2019). ABI score ≤ 0.4 is classified as severe PAD, 0.4-6.9 is classified as moderate PAD, 7.0-0.9 is classified as mild PAD, 0.9-1.3 is classified as normal, and >1.3 is classified as non-compressible vessels (34).

Diabetic peripheral neuropathy is length-dependent sensorimotor polyneuropathy attributable to metabolic and microvessel alterations as a result of chronic hyperglycemia exposure and cardiovascular risk covariates (35). Peripheral neuropathy consists of motoric neuropathy, sensory neuropathy, and autonomic neuropathy. Motoric neuropathy is evaluated visually by the presence of changes in the shape of the fingers and/or soles of the feet, muscle atrophy, and bone protrusions. Sensory neuropathy is examined by subjective and objective measures. Subjective measure is obtained by asking if there are varying pain sensations, such as numbness, tingling, and impaired sensation to light touch. Objective measure is obtained by light touch, monofilament test, and vibration sensation test. Light touch and monofilament test are conducted at 3 points, which are great toes, plantar side of first metatarsophalangeal, and plantar side of fifth metatarsophalangeal. Monofilament test is conducted using 10-G Semmes-Weinstein. Vibration sensation test is conducted using 128 Hz tuning fork, placed on the tip of the great toe. Autonomic neuropathy is evaluated visually by the presence of dry scaly skin or cracked skin.

#### Baseline visit laboratory measures

Fasting blood glucose (FBG) will be measured using blood glucometer (Accu-Chek Performa, Roche Diabetes Care Inc, Meylan, France) applied in capillary blood. Peripheral blood is collected in EDTA and SST Vacutainers (BD, Franklin Lakes, New Jersey, USA). Midstream urine is collected in urine container (OneMed Health Care, Indonesia). HbA1C will be measured using 2 methods: reversed-phase high performance liquid chromatography (HPLC) as the standard method and capillary electrophoresis as comparison using point-of-care testing (POCT). Venous blood sample is analyzed based on HPLC method using HA-8180V analyzer (Arkray Inc., Koji, Konan-Cho, Koka Shi, Shiga, Japan), while capillary blood sample is analyzed based on POCT using TheLab001 analyzer (Arkray Inc., Koji, Konan-Cho, Koka Shi, Shiga, Japan). Total cholesterol, LDL level, HDL level, triglyceride level, estimated glomerular filtration rate (eGFR), and urinary albumin-creatinine ratio (UACR) are measured in an internationally accredited laboratory.

#### Follow-up examination

Follow-up examinations will be conducted 3 months and 6 months after intervention. At 3 months after intervention, subjects are given the second and third questionnaires, evaluating subjective complaints suggestive of T2DM complications, 3-days food recall, sexual activity, family support, global physical activity, hypoglycemia awareness, and global quality of life. Objective examinations include general physical examination and foot examination. FBG, HbA1C, total cholesterol, LDL level, HDL level, triglyceride level, eGFR, and UACR are measured in the same laboratory during baseline visit.

At 6 months after intervention, subjects will again be given the second and third questionnaires, evaluating subjective complaints suggestive of T2DM complications, 3-days food recall, sexual activity, family support, global physical activity, hypoglycemia awareness, and global quality of life. Objective examinations include general physical examination, cardiac examination, and foot examination. FBG, HbA1C, total cholesterol, LDL level, HDL level, triglyceride level, eGFR, and UACR are measured in the same laboratory during baseline visit.

#### Ethics

Ethical clearance is given by The Health Research Ethical Committee, Faculty of Medicine, University of Indonesia with approval number: KET-1146/UN2.F1/ETIK/PPM.00.02. This study is registered as a clinical trial in http://www.clinicalstrials.gov with reference number NCT05090488. Official permission is given by Provincial Health Department of Jakarta to involve PHCs in Jakarta. Prior to the study, subjects will be given informed consent regarding to the study benefits and risks. Subjects have the right to withdraw from the study at any time, without any consequences. All personal information will be kept confidential.

## Discussion

In a previous study conducted in Indonesia, 67.9% T2DM patients had poor glycemic control despite of pharmacological treatment (36). This phenomenon showed that management of T2DM could not merely depend on anti-diabetic drug. Without proper change of daily habit, usage of anti-diabetic drug would be ineffective. An alternative method offered by WHO to promote healthy lifestyle in T2DM patients is through DSME/S. DSMS is a collaborative model in which coaches support their patients with knowledge, skill, and self-awareness so that patients could independently manage their health (37). Due to any limitation during COVID-19 pandemic, the modification of DSMES mode of delivery should be improved in order to overcome the barriers. This study is an open label randomized-controlled trial aim to investigate the role of hybrid model of DSMES in patients with T2DM at primary health care during COVID-19 pandemic.

## Data Availability

All data produced in the present study are available upon reasonable request to the authors

## Abbreviations

T2DM: type-2 diabetes mellitus
PHC: primary health care
OAD: oral anti-diabetic
DSMES: diabetes self-management education and support
SMBG: self-monitoring of blood glucose

## Authors’ contributions

Conceptualization: EY, Syahidatul W, DLT

Data curation: FK, MR, AW, TP

Formal analysis: Syahidatul W, FK, MR, AW, NA, TP

Funding acquisition: EY, DLT, TJET

Investigation: DSH, Suharko S, BD, DP, WW

Methodology: Syahidatul W, LR, YDL, PN

Project administration: APN, AIK, Simon S, GAA, AM, Syahidatul W, MR, AW, NSW, FD, VV

Resources: FK, MR, AW

Software: FK, MR, AW, NA, TP

Supervision: TJET, Sarwono W, IS, IA, DOTLH, ESW, RS, PS

Validation: TJET, Sarwono W, IS, IA, DOTLH, ESW, RS, PS

Visualization: Syahidatul W, EVC

Writing – original draft: Syahidatul W, EVC

Writing – review & editing: EY, Syahidatul W, DLT, LR, YDL, PN, APN, AIK, TJET, Simon S, GAA, AM, Sarwono W, IM, DSH, Suharko S, BD, DP, WW, FK, MR, AR, EVC, TP, ESW, NSW, FD, VV, IA, DOTLH, RS, PS

## Acknowledgement

We would like to thank Vanaya Health Coaching Institute who helped us with coaching training and certification. We also would like to thank all health workers from 10 PHCs who contributed their time for this study: Theresia Indrayanti, Metsy Wendhiani, Ervina Martha, Odilia Ratna, Berlian Situmeang, Hendi Prasetyo, Wiji Ika, Deby Indriati, Mega, Maryam Nur Rahim, Rafida Mardhatila, Shinta Prastyanti, Adriana Eunike, Lily Parliani, Alya Chairunnisa, Almira Sitompul, Syelfia Fitriyani, Imas Yulianti, Mutiara Alderisa, Sudadi, Ajeng Dwiyanti, Sri Wati, Arum Maharani, Dea Noviana, Dhea Nuriza Annisfi, Wasti Evelt Delano, Akhmad Muhajir, Stefany Grandinata Soeseno, Spetiawan Hani Saputro, and Rindiantika. Last but not least, we would like to thank Anindita Wicitra and Noerachma from Anindita Wicitra and Noerachma from Department of Ophthalmology, Faculty of Medicine, Universitas Indonesia/Cipto Mangunkusumo National General Hospital, Jakarta, Indonesia.

